# CLINICAL SAMPLING OF SMALL INTESTINE LUMINAL CONTENT FOR MICROBIOME MULTI-OMICS ANALYSIS: A PERFORMANCE ANALYSIS OF THE SMALL INTESTINE MICROBIOME ASPIRATION (SIMBA) CAPSULE AND BENCHMARKING AGAINST ENDOSCOPY

**DOI:** 10.1101/2024.04.04.24305299

**Authors:** Gang Wang, Cedoljub Bundalovic-Torma, Sharanya Menon, Sabina Bruehlmann, Lynn Wilsack, Renata Rehak, Sahar Bagheri, Mohammad M. Banoei, Lawrence Lou, Yasmin Nasser, Matthew Woo, Ian Lewis, Kathy D. McCoy, Christopher N. Andrews

## Abstract

**Objective:** The small intestine (SI) microbiome is increasingly implicated in both functional gastrointestinal (GI) disorders and a wide range of systemic diseases. However, owing to limitations of traditional GI sampling approaches, the SI remains challenging to directly access on a large scale. This work presents the Small Intestinal MicroBiome Aspiration Capsule (SIMBA) as an effective means for sampling SI luminal content.

**Design:** In an observational clinical study, SIMBA capsules were ingested by both healthy individuals and Irritable Bowel Syndrome (IBS) patients on two successive visits. On a first visit, X-ray scans were used to evaluate SI targeting accuracy. On a second visit, SIMBA capsule ingestion was paired with duodenal endoscopy and saliva samples for reference. For both visits, SIMBA capsules were retrieved with matching fecal samples to evaluate effective sealing during GI transit.

**Results:** X-ray monitoring confirmed all capsules sampled from the distal small intestine with a few (5 of 49) sealing in the proximal colon. Overall, 94% of capsules were retrieved by subjects with median total gut transit time of 47 hours (IQR 24-54). Capsule sampling location and duration was also not significantly affected by IBS. Multi-omics analysis showed that microbiota and metabolomic composition of SIMBA capsules were significantly different to fecal samples, and similar to endoscopic aspirate and cytological brush sampling.

**Conclusions:** The SIMBA capsule reliably captures and preserves SI luminal fluid in a clinically relevant context that is suitable for multi-omics data analysis, comparable to duodenal aspirate, and complements fecal sampling in its broad applicability of use.

**KEY MESSAGES:** **What is already known on this topic**

- Research into the gastrointestinal (GI) microbiome and its role in health and disease is almost completely biased towards the colon due to the reliance on fecal sampling.
- Owing to the lack of reliable and scalable sampling approaches, our knowledge of the small intestine (SI) microbiome is significantly lagging by comparison.

**What this study adds**

- This study presents the Small Intestine Microbiome Aspiration (SIMBA) capsule which targets the distal SI in a reliable and reproducible fashion and collects high-quality multi-omics datasets that are on par with “gold-standard” endoscopic sampling.
- The SIMBA capsule was compared against established sampling methodologies, providing a multi-omics glimpse into the entire biogeographic diversity of the GI tract, revealing substantial and biologically meaningful differences in both microbiome and metabolic profiles that reinforce the significant difference between SI and feces.
- Overall, the SIMBA capsule demonstrates clear and reproducible differences in microbiome composition of the SI that is otherwise lacking from traditionally used fecal sampling.

**How this study might affect research, practice or policy**

- The SIMBA capsule collects high-quality multi-omics datasets that will enable significant insights into the SI microbiome function in health and disease and is ideal for use in research, large-scale clinical or population studies, and diagnostic applications.

## INTRODUCTION

Gut microbiota dysbiosis has been shown to influence the course of not only digestive diseases, but other conditions such as cardiovascular and metabolic diseases, and cancers [1]. The small intestinal microbiome and metabolites of the digestive tract can provide meaningful biological information associated with an individual’s health status but has been little studied in a systematic fashion due to limitations in traditionally used gastrointestinal (GI) sampling approaches, which poses a significant hindrance in our understanding and development of effective diagnostics and treatments of GI-associated diseases.

Currently, there are three main approaches for evaluating the status of the intestinal microenvironment, namely breath hydrogen testing, fecal microbial examination, and invasive endoscopy for digestive sample acquisition [2,3]. However, these approaches have significant disadvantages, including an indirect representation of microbiota and variable reliability with breath testing; non-representation of specific intestinal regions and limited temporal resolution by fecal sample examination; and the invasiveness, inconvenience, and cost of endoscopy. Therefore, the distal small intestine presently remains a ‘black box’ with a total length of 2–5 meters and tortuous anatomy that cannot be easily examined by gastroscopy or standard colonoscopy [4].

There is a critical need for minimally invasive and cost-effective devices that allow direct high-quality sampling in the small intestine. Over the past few years, a few capsule-based sampling systems have been introduced to target the small intestine using passive sampling technologies [5–7]. While early studies in humans have demonstrated luminal fluid collection with multi-omics profiles differing from stool, reference data regarding SI localization and endoscopic sampling has been lacking. To date, there is little data available to evaluate the overall sample quality of these technologies, particularly regarding effectiveness of sealing off in the SI (i.e. avoiding colonic contamination) and sample preservation during passage, collection, and transport for analysis.

The Small Intestine MicroBiome Aspiration (SIMBA) capsule is a pH-based autonomous sampling capsule designed to open and close while transiting the small intestine [7]. The SIMBA capsule has a pH-dependent coating, which keeps it sterile through the oral and gastric regions, and then sloughs off; large sampling ports for content collection of varying consistencies in the SI; effective timed non-electronic sealing performance to avoid colonic contamination; and embedded microbial DNA preserving agents to maintain sample quality during the capsule passage and collection process. The SIMBA capsule was previously shown to safely transit the gastrointestinal tract and collect intestinal fluid samples in real time following probiotic ingestion [7]. This present study aims to demonstrate the sampling quality and efficacy of the SIMBA capsules in both healthy patients and those with altered gut motility with replicate samples at two time points, X-Ray tracking of SI targeting, and direct comparison with endoscopy aspirates and brushings, in addition to saliva and fecal samples for reference. Further, this is the first reported use of a passive capsule sampling technology to investigate the SI environment with spatial and temporal accuracy in humans.

## MATERIALS AND METHODS

### Study Overview

For this study, individuals with previously diagnosed IBS (Rome 4 criteria, diagnosed by a gastroenterologist) and healthy/control volunteers were recruited from clinical practice, recruitment posters at the University of Calgary-affiliated gastroenterology clinics, or through online participation portals. Recruited participants were between 18 – 70 years of age, and selected based on the following criteria; 1) did not have prior gastrointestinal disease, surgery, or radiation treatment; 2) did not use any medications a week prior to the study that would affect GI motility or acidity; 3) if female, were not pregnant, not breastfeeding, and practicing birth control; 4) did not take antibiotics, colon cleanses/colonics, or bowel preparation for colonoscopy within 2 weeks prior to recruitment; 5) if in the control group, did not have fewer than 2 bowel movements a week. The protocol was approved by the University of Calgary’s research ethics board (REB19-0957).

All participants ingested two capsules at two separate visits separated by 7 to 21 days. In the first visit, the two capsules were ingested with the participants in a fasted state (minimum 8 hour fast) with water. For this study a small radiopaque bead was inserted between the external coating and the capsule; when the bead became detached from the capsule on X-ray it indicated that the external coating had sloughed off and that the capsule had started collecting content. SIMBA capsules have a mechanical closure mechanism which can be assessed by X-ray to be open (i.e. collecting content) or closed (i.e. sealed off). X-ray monitoring was performed at regular intervals (approximately every 15 - 45 minutes until capsules completed sampling) to document sample collection start-, end-point locations, sampling durations and timing from ingestion to start- and end-point sampling. After completing their X-ray visit, participants returned home with instructions and materials for retrieving the capsules by screening each bowel movement until they were retrieved. When a capsule was found, subjects also collected a separate fecal sample from the same stool.

Between 7 and 21 days following the initial X-ray visit, fasting participants underwent an esophagogastroduodenoscopy (EGD) procedure to collect a duodenal aspirate, duodenal cytological brush, and saliva sample. Saliva was collected prior to the endoscopy. Standard conscious sedation with fentanyl and midazolam was used, and oral spray anesthetic was not used. The gastroscope was intubated as far as possible into the duodenum (typically to the fourth part; at least third part in all cases). Aspirate was taken first using sterile technique around scope handling and particularly regarding the scope biopsy channel. A sterile aspiration catheter (product number 2181, Hobbs Medical, Inc, Stafford Springs, CT, USA) was used to collect a fluid sample from the distal duodenum. Duodenal mucosal brushing was then taken. All EGD evaluations were visually normal.

The day after endoscopy, two further SIMBA capsules were ingested by the participants. Identical with the first phase of the study, capsules were ingested with water on a fasted stomach and collected along with matched fecal samples at home. Collected capsules and fecal samples were returned via courier immediately and processed within 24 hours upon receipt using sterile technique.

Samples were received, extracted, and prepared for 16S sequencing (International Microbiome Centre, University of Calgary) and metabolomics analysis (Calgary Metabolomics Research Facility, University of Calgary), and analyzed according to protocols described in **Supplementary Methods**.

## RESULTS

The SIMBA capsule technology was evaluated for its ability to reliably and reproducibly collect small intestinal luminal samples by physical assessment as well as by comparison of 16S rRNA gene amplicon sequencing and metabolomics data with samples collected from patient matched feces and endoscopy aspirate. A total of 30 participants were recruited, comprising 10 reference healthy controls and 20 with impaired GI motility (8 IBS-Constipation, 10 IBS-Diarrhea, and 2 IBS-Mixed), with a median age 43 years (min 23; max 67; IQR 32 to 53), and male / female ratio of 12 / 18. Of the 120 total capsule ingestions performed, all capsules were confirmed to have passed spontaneously, with 93% (112 / 120) being successfully retrieved and returned (58 / 60 following X-Ray and 54 / 60 following Endoscopy). Minimal adverse events were reported and were not device-related: 1 event of prolonged capsule retention (> 7 days) in an IBS patient likely associated with opioid used at endoscopy.

### X-Ray Tracking Profiling Demonstrates That the SIMBA Capsule Effectively Targets the Small Intestine and Preserves a High-Quality Sample That is Significantly Distinct From Feces

X-ray tracking of capsule sampling endpoints indicated a high targeting accuracy of the SIMBA capsule to the SI (**Figure 1**). X-rays were read by an expert radiologist blinded to any other subject information other than time post-ingestion. With an X-ray sampling interval of 15-45 minutes, 41 of the 60 capsules ingested had confirmed sampling start locations (start was observed by the marker displacement floating away once the capsule shell dissolved) and 47 had confirmed sampling end locations (end being observation of spring deployment). Sampling location was deemed determinate if it could be confidently assigned by the radiologist and if capsule opening or closing events were seen at least one observation interval after capsule GI-regional transit events. All 41 determinately tracked capsules for start-point initiated sampling in the small intestine, which also included 3 / 4 capsules that were observed to remain in the epigastric region for longer than 2 hours, and of these, one capsule remained in the epigastric region without observed shell dissolution, indicating no impact of gastric pH on SIMBA outer shell integrity. Of the determinate tracked capsules for capsule closure, 44 / 49 (∼90 %) were observed to complete sampling in the SI region, with 15 sampling in the jejunum SI region and 29 in the ileum region. No significant difference in sampling end location was observed across participant motility groups (Chi-squared goodness of fit test p-value ∼ 0.41). The remaining 5 sampled primarily in the distal SI but final sealing was observed in the proximal colon. The median capsule sampling duration from sampling start to end was ∼1.5 hours (min = 1.04 hours; max = 2.02 hours) and was also not found to significantly differ by sampling end location (SI vs. Colon) or participant motility group (Kruskal-Wallis rank-sum test p-value ∼0.78) (**Figure 1B**). The median capsule total GI transit time was ∼46.1 hours and was only found to significantly differ between control and IBS-C groups (median transit time control vs. IBS-C = 29.7 hours vs. 54.7 hours; Mann-Whitney test p-value ∼ 0.006). Excluding one participant where dual-capsules remained in the gastric region for the duration of X-ray observation, X-ray tracking was able to definitively discern that the SIMBA capsule successfully collected a SI sample, with either a determinate starting or end location in the SI, for 29 / 29 ∼ 100% of study participants.

**Figure 1:**
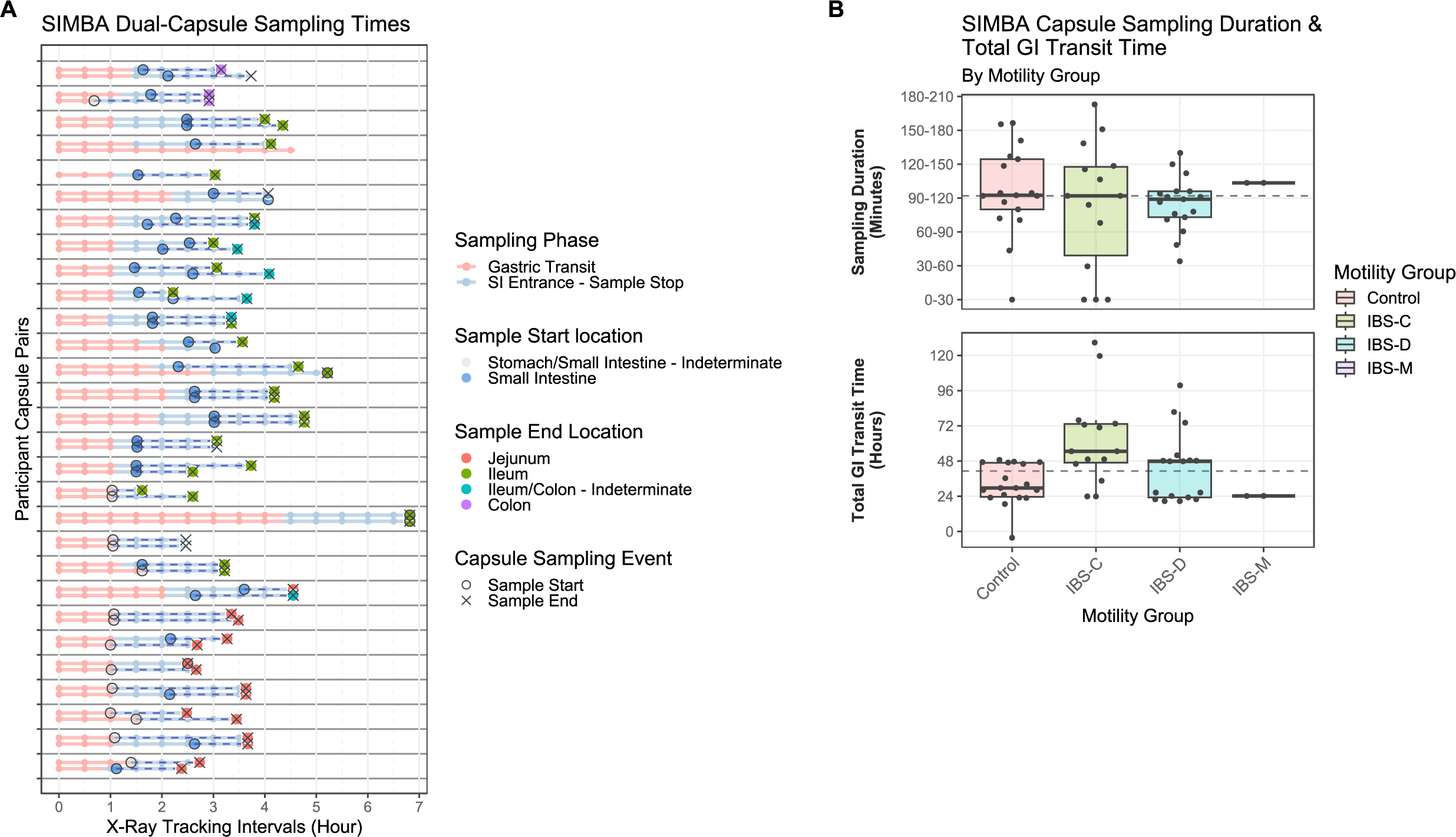
SIMBA capsule sampling performance tracked by X-Ray monitoring. **(A)** Timing of SIMBA capsule sampling events for dual SIMBA capsule ingestions across 30 participants (N = 53 / 60 capsules total). Key events are indicated by line colour (stomach transit, SI entrance to sampling duration), with sampling start (event = outer-shell dissolution) and stop (event = spring deployment & sampling port sealing) indicated by ‘o’ and ‘x’, respectively. Observed capsule sampling start and end locations are indicated by filled point colour. X-axis indicates discrete observation intervals. **(B)** Summary boxplots of corresponding SIMBA capsule sampling durations (top – in X-ray observation intervals of 30 minutes) and total GI transit times (bottom) grouped by participant GI motility status (N = 51 capsules with observed sample endpoints). Dashed lines indicate overall median capsule sampling duration and transit time, respectively.

To evaluate the efficacy of the SIMBA capsule in capturing and preserving an uncontaminated sample of the SI microbiota, microbiome and metabolomics profiles of key GI metabolites (BAs and SCFAs) were compared between SIMBA capsules and matched fecal samples (**Figure 2**). An examination of 16S microbiome taxa plots revealed distinct microbiome compositions between SIMBA capsule and fecal samples, with the former dominated by bacteria of the genera *Streptococcus*, and the latter dominated a more diverse profile of *Blautia, Bacteroidetes*, and *Fecalibacterium,* consistent with previous findings [4]. Principal component analysis of microbiome beta-diversities (weighted Unifrac distance) confirmed a significant difference in microbiome composition between SIMBA capsule and fecal samples (PERMANOVA P-value ∼ 0.001, n = 66 samples) (**Figure 2B**). A number of SIMBA capsule samples were found to have relatively lower diversity microbiome communities as the result of lower total 16S sequencing depth than expected (22 / 58 capsules, corresponding to 1000 reads). Further investigation determined that the most likely cause was the choice of a suboptimal primer for PCR amplification, as a previous study using a primer spanning the V3-V4 region generated an overall 10-fold increase in read depth (i.e. a minimum depth of 3 x 10^4^ total reads across 80 capsule samples) [7]. No significant difference in microbiome composition was observed between capsules which finished sampling in the jejunum vs. Ileum vs. colon (PERMANOVA p-value ∼ 0.344, R^2^ ∼ 0.1) nor by participant IBS status (PERMANOVA p-value ∼ 0.2, R^2^ ∼ 0.13), and did not substantially change with the inclusion of samples with lower sequencing depth.

**Figure 2:**
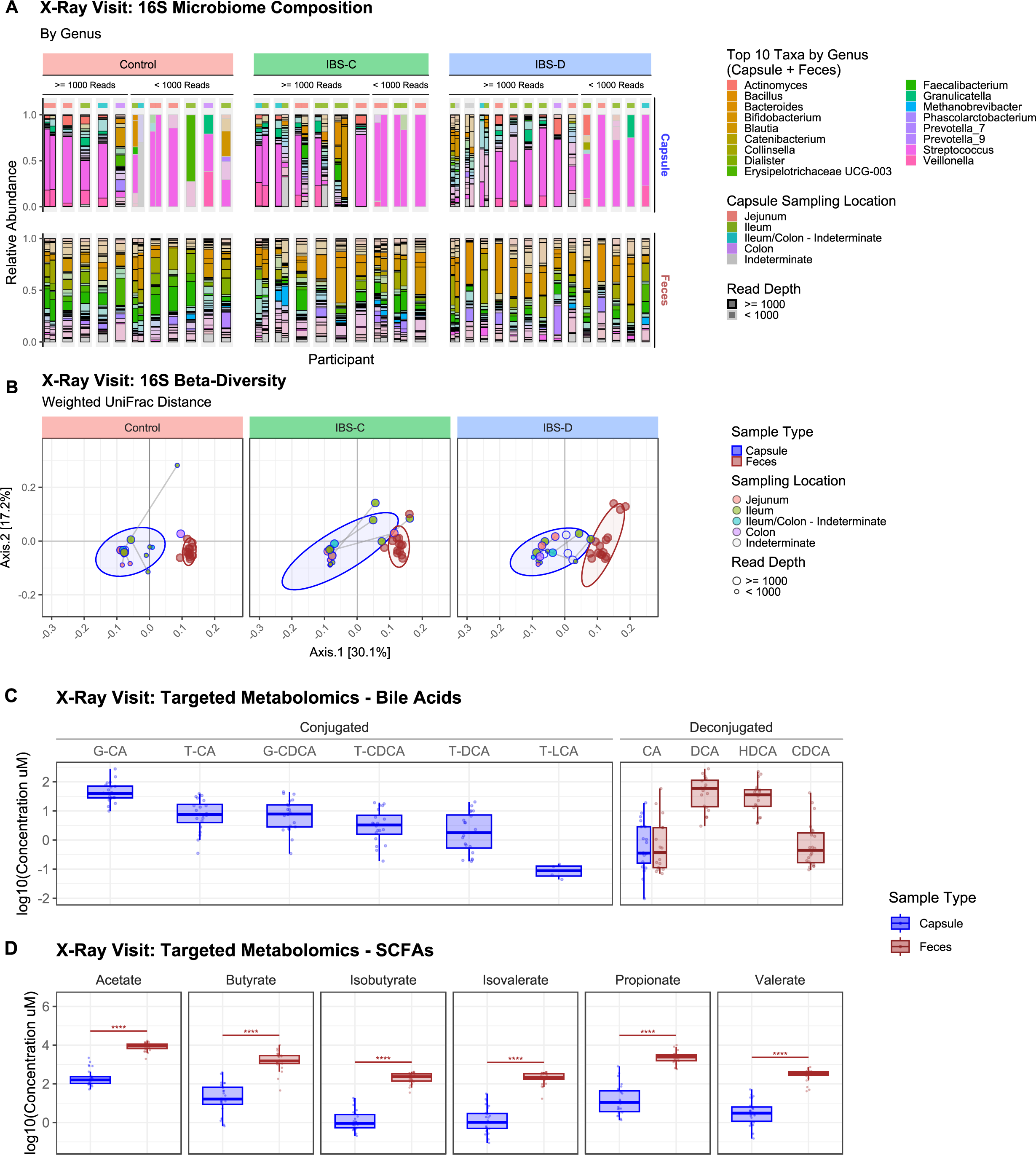
SIMBA Capsule 16S Microbiome Profiles Compared Across Sample Types and Motility Groups and Targeted Metabolomics - X-Ray Visit. **(A)** Taxonomic profiles of bacterial genera identified in SIMBA capsule and fecal samples distinguished by patient motility group. Top row - SIMBA capsules with corresponding X-ray sampling endpoint GI locations indicated above taxa barplots, distinguished by higher (>= 1000 total reads – black outline) and lower (< 1000 total reads – grey outline) sequencing depth. Bottom row - Corresponding matched fecal samples (if available). Top-10 taxa by relative abundance in SIMBA capsules and fecal samples are indicated by opaque filled bars. **(B)** Principal co-ordinates analysis ordination plot of sample beta-diversity (weighted UniFrac distances of ASV relative abundances), with ellipses drawn around samples belonging to the same sample type, points coloured by SIMBA capsule sampling location, and lines joining dual capsules ingested by the same participants. Points are sized according to sequencing depth (>= 1000 vs. < 1000 total reads). **(C)** Targeted metabolomics of conjugated and deconjugated bile-acid absolute concentrations across sample types. Bile acid acronyms are as follows: Conjugated – taurine and/or glycine conjugated forms of cholic acid (T/G-CA), chenodeoxychoic acid (T/C-CDCA), deoxycholic acid (T-DCA) and lithocholic acid (T-LCA); Deconjugated – DCA, CDCA, and hyodeoxycholic acid (HDCA). **(D)** Targeted metabolomics of short-chain fatty acid absolute concentrations across sample types, with FDR adjusted Kruskal-Wallis p-values indicated above pairwise sample type comparisons, and coloured according to sample type with higher median concentration.

The differences between SIMBA capsule and fecal sampling were also shown by the concentrations of key GI metabolites. For example, the concentrations of all SCFAs examined were found to be significantly increased in fecal samples compared to SIMBA capsules (Mann-Whitney test p-value range: 1.65 x10^14^ – 2.29 x 10^12^) (**Figure 2C**). Furthermore, a striking difference was observed between conjugated and deconjugated BAs, which were nearly all exclusively associated with SIMBA capsule and fecal samples, respectively (**Figure 2D**). The only exception was the presence of the primary cholic acid (CA) in feces, which was most likely the result of production via alternate colonic microbial deconjugation pathways [8]. Together these results confirm that SIMBA capsules capture a distinct microbiome and metabolomic profile from the SI that is effectively preserved against fecal contamination during GI transit and performs robustly under different gut motility conditions.

### SIMBA Capsules Effectively Sample the SI On Par with Gold-Standard Endoscopy As Corroborated By Key Differences in Microbiome and Metabolomics Profiles Compared to Feces

As a final evaluation of SIMBA capsule performance as an accurate and reliable SI sampling tool, microbiome composition (using 16S rRNA gene sequencing) and metabolomic profiles from a second round of SIMBA capsule ingestions were compared against gold-standard endoscopic aspirate and cytology brush samples from the duodenum (**Figure 3**). In addition to fecal samples, which served to demonstrate the effective sealing performance of SIMBA capsules against potential fecal contamination, saliva samples were also included to assess potential oral contamination of endoscopy sampling.

**Figure 3:**
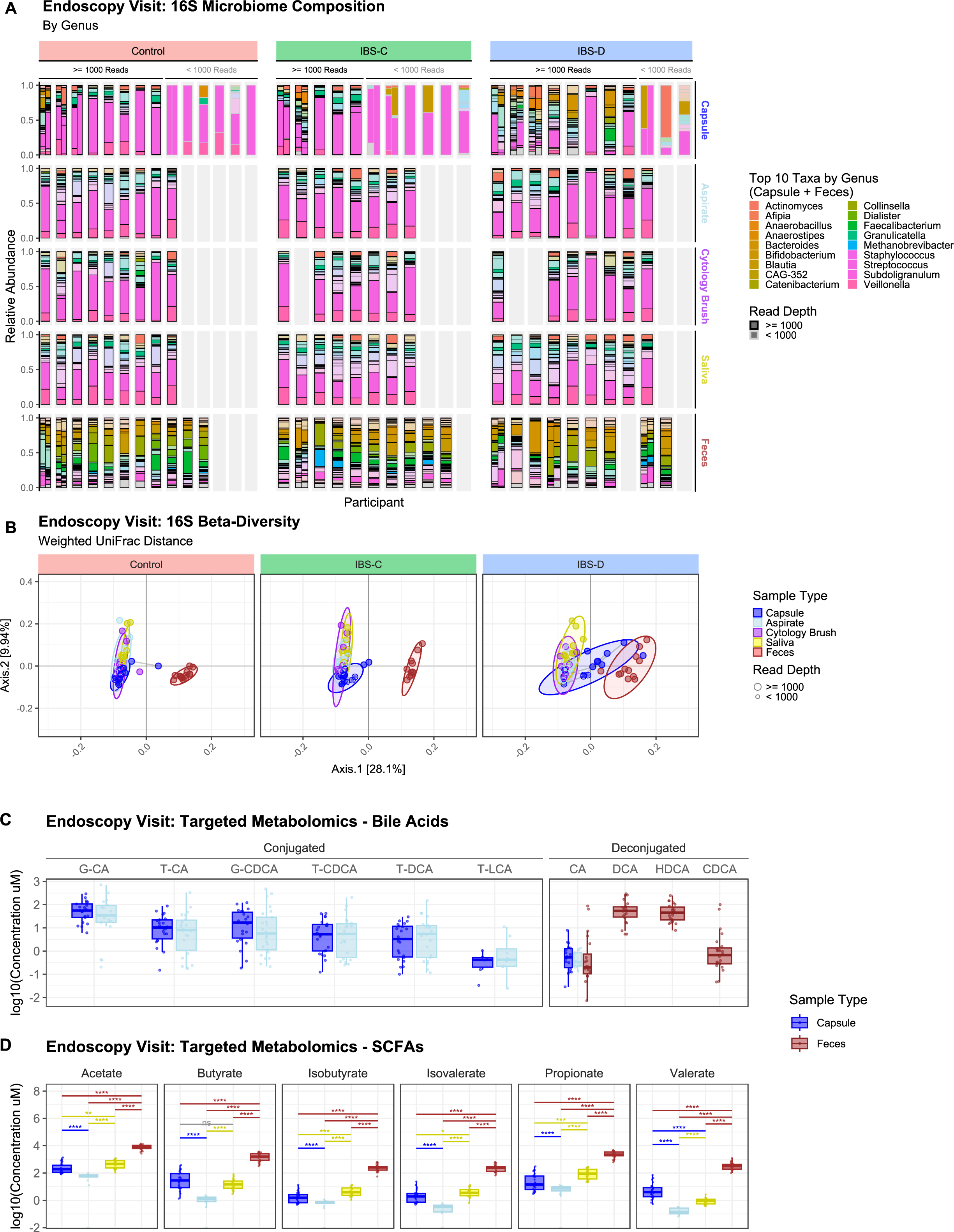
SIMBA Capsule 16S Microbiome Profiles Compared Across Sample Types and Motility Groups and Targeted Metabolomics - Endoscopy Visit. **(A)** Taxonomic profiles of bacterial genera identified in SIMBA capsules (top row) compared to matched endoscopic aspirate, cytological brush, saliva, and feces samples distinguished by participant motility group. Top Facet - SIMBA capsule samples arranged by participant into groups by higher and lower sequencing depth, >= 1000 total reads – black outline, < 1000 total reads – grey outline, respectively. Lower Facets - Corresponding samples collected during endoscopy visit, and matched fecal samples. Top-10 taxa by relative abundance in SIMBA capsules or fecal samples are indicated by opaque filled bars. Note: ‘missing’ fecal samples correspond to capsule pairs collected in a single bowel movement. **(B)** Principal co-ordinates analysis ordination plot of sample beta-diversity (weighted UniFrac distances of ASV relative abundances), with ellipses and points coloured by SIMBA capsule sample type, and lines joining dual capsules ingested by the same participants. Points are sized according to sequencing depth (>= 1000 vs. < 1000 total reads). **(C)** Targeted metabolomics of conjugated and degonjugated bile-acid absolute concentrations across sample types. **(D)** Short-Chain Fatty Acid absolute concentrations across sample types with and significant pairwise differences (FDR adjusted Kruskal-Wallis test p-values < 0.05) indicated and above and coloured by sample type with greater median log10 concentration. For definition of bile acid acronyms see the caption of Figure 2.

Principal co-ordinates analysis of 16S microbiome beta-diversities revealed a noticeable degree of overlap between SIMBA capsule, endoscopic aspirate and cytological brush samples, particularly on the primary-axis of variation (∼28.1% total variance explained) (**Figure 3B**). Interestingly, the second axis of variation (∼9.9% of total variance explained) also indicated a separation between SIMBA capsules from a subset of endoscopic and saliva samples, likely reflecting biologically relevant differences between duodenal and ileum microbiome communities resulting from underlying physiological pH gradients (see following section). Further PERMANOVA statistical testing (regressing weighted Unifrac beta diversity against sample type as an ordered factor with SIMBA capsules as the intercept) did reveal a significant difference in 16S microbial composition between SIMBA capsules and other sampling methods (p-value ∼ 0.001, R^2^ ∼ 0.41). However when broken down by sample type the majority of variation was contributed by fecal and saliva samples (feces PERMANOVA p-value ∼ 0.001, R^2^ ∼ 0.17; saliva PERMANOVA p-value ∼ 0.001,S R^2^ ∼ 0.13), while endoscopic aspirate and cytology brush contributed a substantially lower effect-size in terms of proportion of variance explained (endoscopic aspirate PERMANOVA p-value ∼ 0.001, R^2^ ∼ 0.048; cytology brush PERMANOVA p-value ∼ 0.001, R^2^ ∼ 0.061). Overall, microbiome profiles captured by capsule and endoscopy were both substantially different to fecal samples and to similar degrees (capsule vs. feces median Unifrac distances ∼ 0.538; endoscopic aspirate vs. feces ∼ 0.554; cytology brush vs. feces ∼ 0.541).

The analysis of targeted metabolomics profiles also demonstrated that both endoscopic aspirates and SIMBA capsules recapitulated significant differences in BA and SCFA concentrations in comparison to fecal samples. The presence of conjugated BAs was also found to be exclusive to the SI, with SIMBA capsules showing no significant differences in concentration compared to gold standard endoscopic aspirates (Mann-Whitney test FDR adjusted p-values: min = 0.194, max = 0.917, mean ∼ 0.5). Deconjugated BAs were again exclusively found in feces, except for cholic acid (CA) which are also detected in the Capsule and Aspirate samples (**Figure 3C**).

SCFA concentrations were also found to be significantly increased in feces in comparison to both SIMBA capsule and endoscopic aspirates (Mann-Whitney test FDR-corrected p-values, endoscopic aspirate vs. feces = 2.72 x 10^-9^ to 1.23 x 10^-8^), similar to previous results from the X-ray visit. Interestingly, SCFA concentrations were also found to be significantly increased in SIMBA capsules relative to endoscopic aspirates (Mann-Whitney test FDR-corrected p-values, SIMBA capsule vs. endoscopic aspirate = 2.72 x 10^-9^ to 3.84 x 10^-6^) (**Figure 3D**). This result suggests an increasing gradient in SCFA production from proximal SI (endoscopy), distal SI (SIMBA capsule), and colon (feces), and furthermore implies that SCFA production is not an exclusive metabolic function of the colonic microbiome.

Additional semi-targeted metabolomics analyses of a panel of 85 metabolites also revealed a strong differentiation between SIMBA capsules and fecal sample and similarity to endoscopic aspirates, as indicated by hierarchical clustering of metabolite profiles (**Figure 4**). Given the substantial differences in total spectral abundance between fecal and SI samples (feces vs. SIMBA capsule and endoscopic aspirate median log10 total spectral abundances: ∼7.5 vs. 8.3), spectral counts were normalized using the following procedure to aid comparison [9,10]: normalization by median metabolite spectral count; log10 transformation; and sample-wise auto-correlation/unit scaling of metabolite profiles. Additional K-means clustering of log10 transformed metabolite intensities further revealed markedly different patterns of intensity for different sets of metabolites across sample types (**Figure 4** – red asterisks and **Supplemental Figure 1A**). Although the majority of metabolites (endoscopy visit) were generally of high intensity in fecal samples and low in saliva (63/85 significantly elevated metabolite concentrations in feces by Mann-Whitney test FDR adjusted p-value <= 0.05), several were identified (10/85) that were significantly increased in SIMBA capsule samples (Mann-Whitney test FDR adjusted p-value range < 0.04 to 1.5 x 10^-9^), of which (8/10) were also significantly increased in endoscopic aspirates (**Supplemental Figure 1B**). As expected from the targeted metabolomics analyses, the primary bile-acid glycocholate was identified, as well as several amino-acids (L-Arginine, L-Histidine, and L-Cysteine). Taken together, these results demonstrate that the SIMBA capsule performs on par with endoscopy in sampling the SI, capturing microbiome profiles and broad metabolic profile distributions that are significantly distinct from feces, and identifying metabolic markers associated with important physiological differences between the SI and colon.

**Figure 4:**
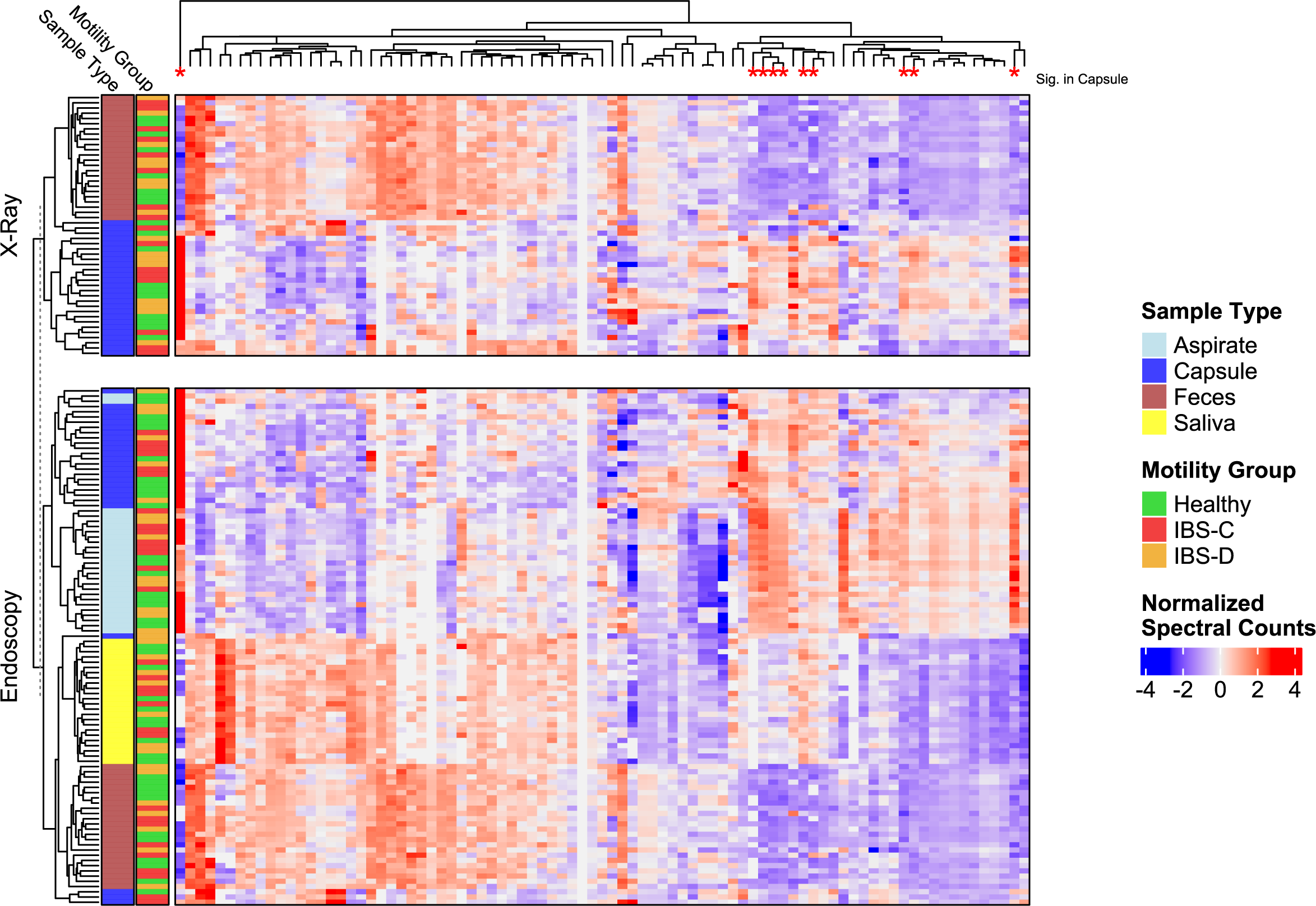
Semi-targeted Metabolomics Profiling Across Sample Types and Visits. Hierarchically clustered heatmap showing normalized spectral intensities for 85 metabolites (columns) across samples (rows) collected during X-Ray and Endoscopy visits. Sample dendrogram indicates distinct clustering of SIMBA capsule and fecal samples across visits, and co-clustering of SIMBA and endoscopic aspirates (bottom). Red asterisks on tips of column dendrogram indicate metabolites with significantly increased log10 spectral intensities in SIMBA capsules vs. feces (Mann-Whitney test FDR adjusted p-value <= 0.05 – see Supplemental Figure 1).

### Comparison of SIMBA Capsule to Endoscopic Aspirates Reveals Subtle Differences in Bacterial Taxonomic Composition Consistent with Differences in SI Sampling Location

To understand whether the differences in 16S microbiome composition between SIMBA capsule and endoscopy samples were due to potential oral contamination, or a biologically meaningful difference in community composition due to differences in SI sampling location, a summary of all unique and shared amplicon sequence variants (ASVs) detected across the distinct sample types was generated, representing the biogeographic distribution of bacteria across the GI tract (**Figure 5**).

**Figure 5:**
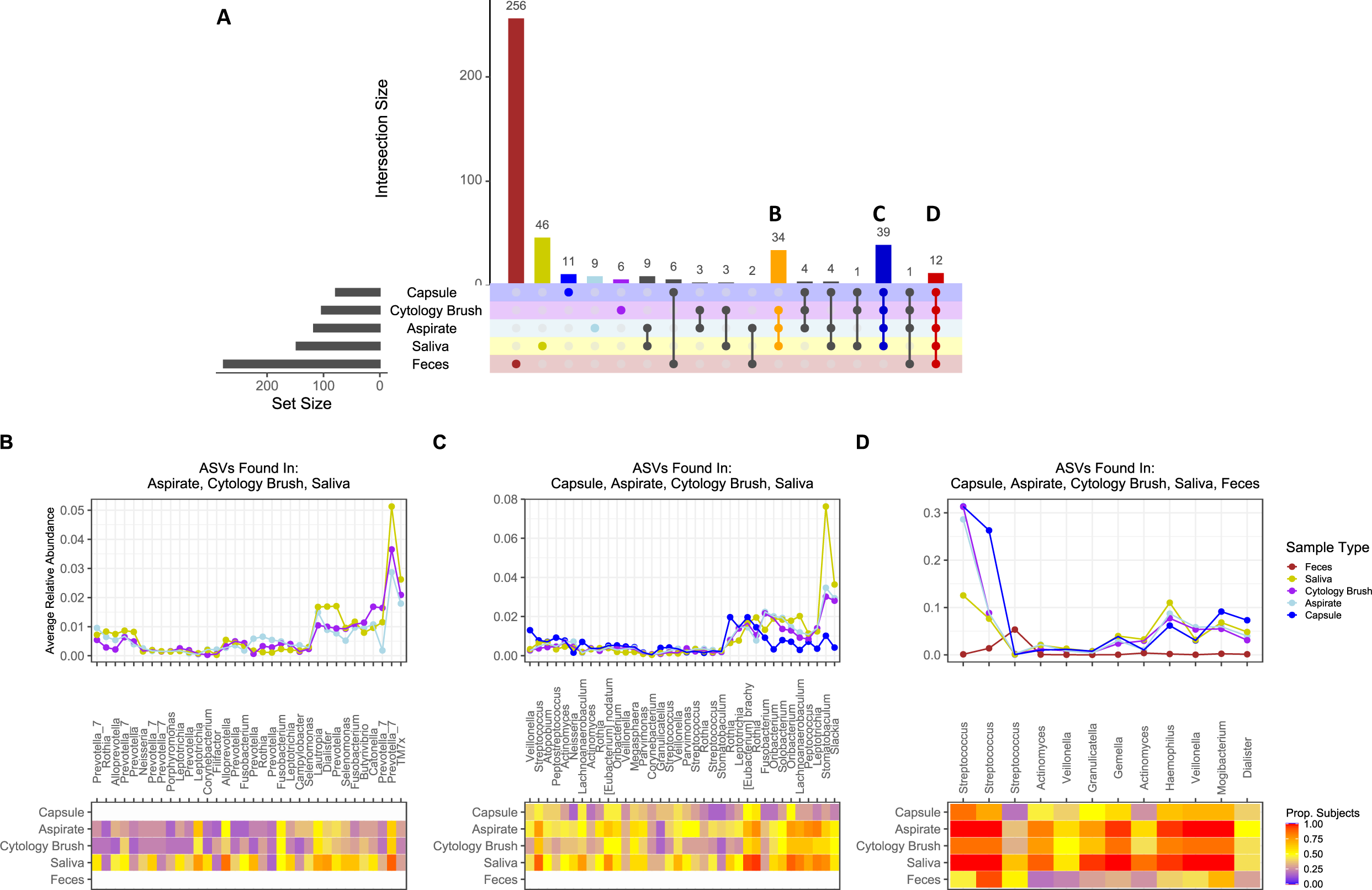
Intersection of Microbial ASVs Identified Across Sample Types. **(A)** Upset plot illustrating the distribution of identified ASVs (total number of participants >= 5 and mean relative abundance >= 1e-5) across sample type intersection sets (number of unique ASVs indicated above bars). Intersections of interest are highlighted and labelled corresponding to the accompanying panels of ASV mean sample relative abundances: **(B)** ASVs identified exclusively between endoscopy and saliva samples indicating proximity / overlap of oral bacteria (genera given as species assignments are largely unavailable) in the duodenal region of the SI; **(C)** ASVs shared between SIMBA capsule, endoscopy and saliva, showing different profiles of SI associated bacteria originating from the oral microbiome; **(D)** All sample types, indicating predominance and enrichment of *Streptococcus* in SIMBA capsules and endoscopic samples.

Although the vast majority of ASVs detected were unique to fecal samples (256/446 ∼ 57% of ASVs), followed by saliva samples (46/446 ∼ 10% of ASVs detected), several intersections of interest were found that revealed insights into the distribution of bacterial genera across the GI tract. The first intersection of particular interest represented ASVs that were only identified in endoscopic aspirate and cytology brush, and saliva samples (34/446 ∼ 7% of ASVs detected) (**Figure 5B**). Notably, this intersection contained several ASVs annotated to the family Prevotellaceae, a known acid-tolerant bacterium present in the oral microbiome [11], which were frequently detected (>= 50%) across saliva and endoscopic samples (duodenum), but notably absent in SIMBA capsules (jejunum + ileum). These results would indicate that the differences between SIMBA capsule and endoscopy are likely reflecting biologically relevant regional differences in microbiome composition influenced by proximity of sampling location to the stomach [4]. A second substantial intersection (**Figure 5C**) included ASVs identified in SIMBA capsule and endoscopy and saliva samples (39/446 ∼ 8%), which were relatively increased in relative abundance and prevalence in endoscopy and saliva compared to SIMBA capsule. The last intersection of interest (**Figure 5D**) contained ASVs identified in all sample types (12 / 446 ∼ 2%), of which *Streptococcus*, a keystone genus of the small intestine [12], was found to be particularly dominant across SIMBA capsule, endoscopic aspirate, and cytological brush samples (∼30% relative abundance) and reduced in saliva (∼10%) and feces (< 1%). Taken together, these results further support that the SIMBA capsule is capturing a sample representative of the small intestine, specifically the distal region.

### SIMBA Capsule Multi-Omics Datasets Are Reproducible Over Time

No significant differences were found between SIMBA capsule 16S profiles between Endoscopy and X-Ray tracking visits regarding the level of between-sample beta-diversity (SIMBA capsule vs. feces Mann-Whitney test p-value ∼ 0.90; weighted Unifrac distance of microbiome proportional abundance profiles) as well within-individual beta-diversity by sample type (Mann-Whitney test p-values: SIMBA capsule X-Ray vs. Endoscopy Visits ∼ 0.219; Feces X-Ray Vs. Endoscopy visits ∼ 0.752) (**Figure 6A**). Similar to 16S microbiome profiles, metabolomics data generated from SIMBA capsules was also found to be highly reproducible between visits, as demonstrated by a semi-untargeted screen of 85 metabolites at the between-sample (SIMBA capsule vs. feces Mann-Whitney test p-value ∼ 0.179; Euclidean distances of log10 transformed metabolite spectral count profiles) and within-individual by sample type (Mann-Whitney test p-values: SIMBA capsule X-Ray vs. Endoscopy Visits ∼ 1.0; Feces X-Ray Vs. Endoscopy visits ∼ 1.0) levels (**Figure 6B**).

**Figure 6:**
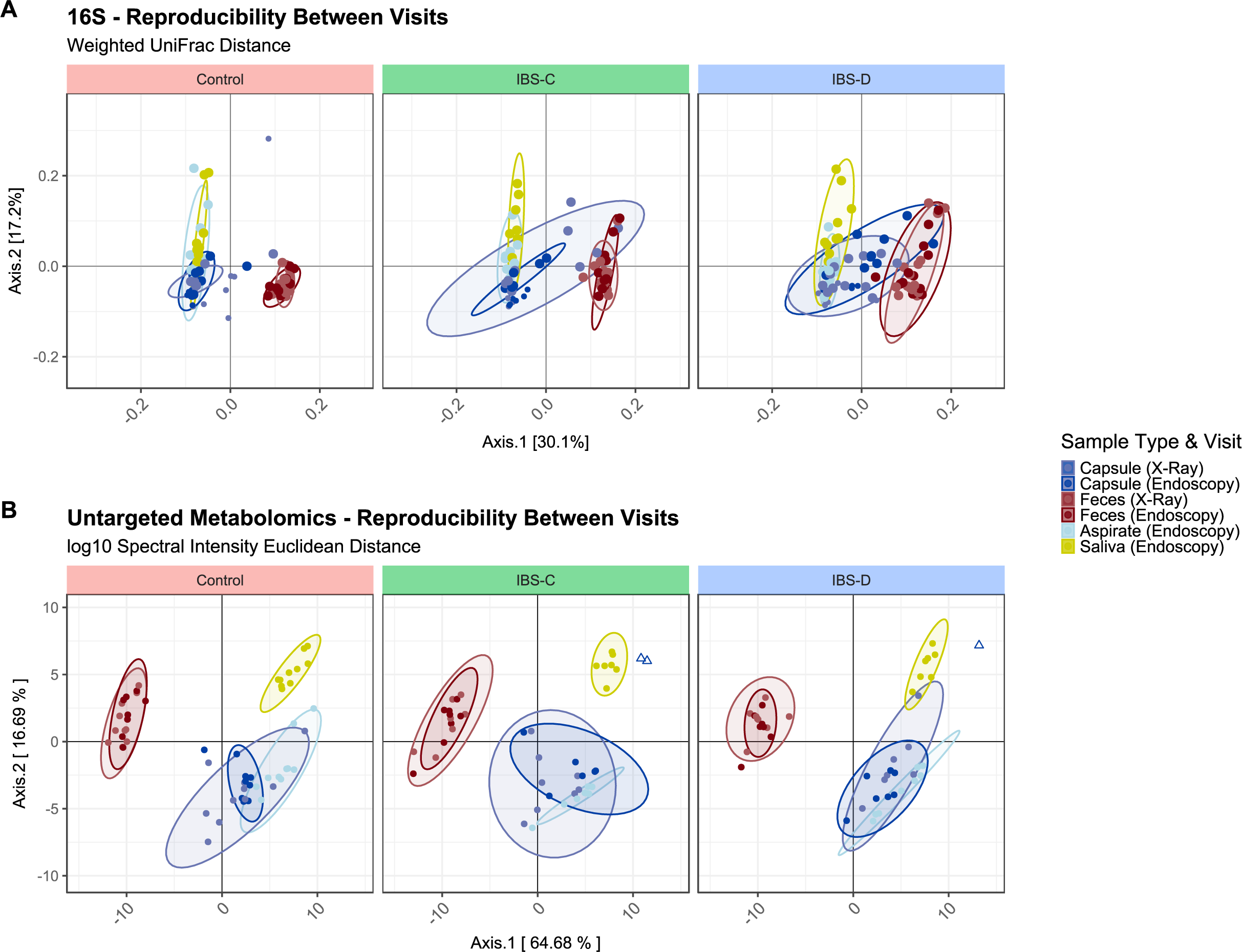
Reproducibility of Multi-Omics Datasets Across X-Ray and Endoscopy Visits: Principal co-ordinate ordination plots of: (A). 16S beta-diversity (weighted Unifrac distanace corresponding to data shown in Figures 3 & 4) and; (B) Untargeted metabolomics profiles (log10 transformed spectral intensities corresponding to data shown in Figure 4) illustrating similarity of SIMBA capsule samples across sample types and visits. Metabolomics samples identified as outliers (< 1.5 – IQR) based on total sample spectral abundance are indicated as triangles and excluded from the calculation of sample type ellipses.

Interestingly, a greater degree of variability in microbiome diversity was previously observed across participant GI motility groups for SIMBA capsule samples, which was not apparent from metabolomics data. These differences were only found to be statistically significant for SIMBA capsules collected from IBS-D participants during the endoscopy visit and were also supported by endoscopic aspirate and cytology brush samples (SIMBA Capsule PERMANOVA p-value ∼ 0.02 and R^2^ ∼ 0.29; endoscopic aspirate PERMANOVA p-value ∼ 0.006, R^2^ ∼ 0.027; endoscopic cytological brush PERMANOVA p-value ∼ 0.014, R^2^ ∼ 0.17 respectively), but were not observed by fecal sampling (IBS-C p-value ∼ 0.17, IBS-D p-value ∼ 0.92). These results may indicate potential indications of increased microbial dysbiosis, i.e. alteration in microbiome composition of resident SI taxa compared to controls [13], in IBS-C participants, but owing to the limitations in power and scope of this present project, no definitive conclusions can be drawn. However, given previous reports of dysbiosis associated with IBS status [3,14,15], these findings may be biologically relevant and would certainly merit further investigation with expanded clinical cohorts.

## DISCUSSION

Sampling methods which enable *a nuanced spatial and longitudinal understanding of the microbiome across its biogeography* [16] will be key towards advancing our knowledge of the role of the GI microbiome, enabling us to investigate how different external factors affect its long-term trajectory and ultimately impact human wellbeing. The SI represents a frontier in our knowledge of the GI tract and has been relatively little explored compared to the colon due to its relative difficulty to access. In addition, the lower biomass of the SI microbiome and its high degree of turnover [12] requires sampling methods that are robust to its inherent variability and provide sufficient temporal resolution. Recent large-scale clinical recruitment cohort studies have spearheaded efforts to address this problem through biopsy or endoscopic aspirate sampling [17,18], however there is still a dearth in our knowledge of the SI microbiome [1,2,4]. Although these approaches represent the gold-standard in direct-sampling of the SI microbiome, they are not feasible, both financially and technically, for broad, large-scale deployment. Novel SI sampling technologies hold great promise to enrich the arsenal of microbiome research [6] and require robust and reliable benchmarking for research and clinical use.

To this end, the SIMBA capsule was developed. Benchmark testing with X-ray tracking data confirmed that SIMBA capsule sampling was localized to the jejunum and ileum of the SI, and 16S microbiome profiles demonstrated that SIMBA capsule samples were significantly distinct from feces, indicating effective sample containment during GI transit. Furthermore, of 5 SIMBA capsules which were observed to complete sampling in the colon, and 9 samples with indeterminate sampling endpoint location, none appeared to have either a ‘fecal-like’ microbiome profile nor the presence of secondary BAs, and thus captured a representative sample of the SI. One additional and key difference of the SIMBA capsule from fecal sampling is its ability to provide a time referenced sample from the GI tract. The average duration of SIMBA capsule sampling lasted roughly ∼ 1.5 hours (+/-0.5 hours), compared to an average total GI transit time of ∼ 48 hours (IQR ∼ 24 – 53 hours) for matched fecal samples. Importantly, sampling duration, start, and end locations did not significantly differ between control and IBS subjects, demonstrating that the SIMBA capsule has a broad applicability in real-world clinical settings and provides a time-stamped and thus more reliable sample of the GI microbiome that is of high practical value for intervention studies.

The combination of consistency in sampling localization and sampling duration is an important factor in minimizing variability and capturing a representative and reproducible sample of the SI. Indeed, SIMBA capsules were effective in collecting SI samples with variability on par with endoscopy, and high reproducibility across study visits (at least 1 week apart) with no significant differences observed in both 16S and metabolomics profiling compared to feces. Compared to a previous study [7], we did note an unexpected increase in SIMBA capsule samples with low total sequencing depth (< 1000 reads), reflected by a decrease in 16S profile microbiome diversity, which was attributed to suboptimal 16S primer selection. For example, previously using a V3-V4 primer for 16S amplicon sequencing resulted in a sequencing depth of 7 x 10^4^ for SIMBA capsule samples, which was roughly equivalent to the sequencing depth obtained from corresponding fecal samples, despite an over 100 average fold difference in DNA concentration. This illustrates the critical role of optimization of sample processing for low-biomass samples, especially for drawing reliable conclusions regarding microbiome compositional differences between sites with substantial differences in biomass.

Despite the well-known challenges associated with sampling low-biomass environments, the SIMBA capsule successfully captured a high-quality sample of the SI amenable to generating high-quality multi-omics datasets and also identified biologically meaningful signals of regional variation in microbial communities throughout the GI tract [17,18]. Comparison of SIMBA capsule bile acid and SCFA concentrations further confirmed that the SIMBA capsule can effectively capture meaningful signals of microbiome functional diversity along the GI tract. These metabolite classes are important hallmarks of the diverse metabolic activities and biological roles of upper and lower GI tract/microbiome, and their disruption is increasingly implicated in a broad range of diseases [19,20]. Therefore, spatially resolved microbiome sampling can yield valuable insights about how to effectively modulate microbiome activity for therapeutic intervention [4]. Expanded semi-target metabolomics analysis of 85 additional metabolites not only further confirmed the similarity of SIMBA capsule endoscopic aspirates, but also revealed substantially different distributions in metabolic profiles compared to fecal samples. Specifically, the majority of metabolites investigated appeared to be significantly increased in feces, therefore reaffirming that fecal samples are inadequate for understanding the metabolic activities of the GI as a whole and the critical importance of sampling methods that can accurately access the SI.

Furthermore, SIMBA sample 16S microbiome profiles were highly similar to endoscopy (duodenal aspirates) in their clear distinction from feces but differed by the absence of specific oral microbial taxa that were present in the latter, notably *Prevotella*, which indicate these differences to likely reflect subtle physiological-relevant differences in sampling location between SIMBA capsules (jejunum/ileum) and endoscopy (duodenum). The presence of Prevotellaceae in the SI has been previously reported [11] and its presence may be the result of increased tolerance the low-pH environment of the duodenal sampling location of endoscopic samples, compared to the jejunum/ileum [16] where our previous X-Ray tracking results indicate that the majority of SIMBA capsules begin and end sampling.

Although clinical diagnosis and further investigation of group-wise differences by IBS groups are hampered by the limited sample size and were beyond the intended aims of this study, we hope this work highlights the significant limitations of fecal sampling in microbiome research, and the promising potential and advantages of the SIMBA capsule as a clinical investigative tool that effectively samples a snapshot of the SI microbiome and generates high-quality multi-omics datasets on par with gold-standard endoscopic aspirate. The ultimate goal of this work is to enable microbiome research with crucial real-time insights derived from multi-omics datasets that provide more than a mere description of dysbiosis but a model of the microbiome that can unlock its therapeutic potential in human wellbeing.

## CONTRIBUTORSHIP STATEMENT

G.W., S.Br., and C.N.A. conceived and planned the experiments. G.W., S.M., L.W., R.R., L.L. and C.N.A carried out the experiments. S.M. contributed to sample preparation. M.M.B. and I.L. generated metabolomics data. S.Ba. contributed preliminary 16S sequencing analyses. G.W., C.B-T., S.Br., L.L., and C.N.A. contributed to the interpretation of the results. C.B-T. contributed additional analyses and generated manuscript figures. G.W and C.B-T. contributed equally to the writing of the manuscript. All authors provided critical feedback and helped shape the research, analysis, and manuscript.

## DATA AVAILIBILITY STATEMENT

Corresponding raw .fastq files for 16S analyses will be deposited with links to BioProject accession number PRJNAxxxxxx in the NCBI BioProject database (https://www.ncbi.nlm.nih.gov/bioproject/) upon peer reviewed publication.

## ETHICS

This study was conducted under the guidance and approval of the University Conjoint Ethics Review Board (REB19-0957) and Investigational Testing Authorization from Health Canada (ITA 310032).

## Data Availability

All data produced in the present study are available upon reasonable request to the authors, and will be made available online in the NCBI BioProject database upon peer reviewed publication.

