## Supplementary Methods for "CLINICAL SAMPLING OF SMALL INTESTINE LUMINAL CONTENT FOR MICROBIOME MULTI-OMICS ANALYSIS: A PERFORMANCE ANALYSIS OF THE SMALL INTESTINE MICROBIOME ASPIRATION (SIMBA) CAPSULE AND BENCHMARKING AGAINST ENDOSCOPY"

#### Sample Removal from the SIMBA Capsule

The intestinal samples were removed from the collected capsules using an aseptic technique that included cleaning and disinfecting the capsules. The liquid and fiber portions were transferred into separate 1.7 ml microcentrifuge tubes after opening the capsule. Liquid samples were centrifuged at 4,000 rpm for 10 minutes at 4°C, 40 µL of the supernatant was removed for metabolite extraction and the remaining liquid sample was returned to the fiber portion for DNA extraction.

The sample weight (mg) was calculated by subtracting the weight of the empty tube from the weight of the tube containing sample. The weight of the fiber (20 mg) was subsequently subtracted from the sample weight calculated above.

### 16S SEQUENCING AND MICROBIOME PROFILING

#### DNA extraction and quantification

*For the samples from the SIMBA capsules*, the liquid and fiber portions were combined in the PowerBead Pro tube provided by QIAamp® PowerFecal® Pro DNA Kit (QIAGEN, Hilden, Germany). DNA extraction was performed following the manufacturer's instructions.

*For saliva, aspirate, and fecal samples*, 250 mg (or µL) of sample was added to the PowerBead Pro tube; DNA extraction was performed following the manufacturer's instruction.

The concentrations of DNA were determined by using the Qubit dsDNA HS Assay kit and the Qubit 4.0 fluorometer following the manufacturer's instructions (Thermo Fisher Scientific, Waltham, MA, USA).

#### 16S rRNA gene sequence analysis

16S rRNA metagenomic sequencing was performed by the International Microbiome Center at the University of Calgary. Whole genomic DNA was extracted from each sample using the PowerFecal® Pro DNA Kit as per the manufacturer's instructions. The V4 variable region of the 16S rRNA gene was amplified using PCR primers with internal barcodes (primers shown below) in a 25 cycle PCR using the KAPA HiFi HotStart master mix (Roche Diagnostics GmbH, Mannheim, Germany). The conditions for the thermocycler were as follows: 98°C for 2 minutes, followed by 25 cycles of 98°C for 30 seconds, 55°C for 30 seconds and 72°C for 20 seconds, after which a final elongation step at 72°C for 7 minutes. Amplified PCR products were checked in a 1% agarose gel. The PCR products were then purified using NucleoMag NGS Clean-up and Size Select (Macherey-Nagel, Dueren, Germany) and concentrations normalized using SequalPrep Normalization Plate (Invitrogen, Grand Island, New York, USA). Amplicons were pooled and concentration and quality were determined using the Qubit HS DNA kit (Invitrogen, Grand Island, New York, USA) and the Tapestation D1000 assay (Agilent Technologies, Santa Clara, CA, USA), respectively. Amplicon sequencing was done on a MiSeq Benchtop DNA sequencer (Illumina, San Diego, CA, USA) using a V2-500 cycle kit (Illumina, San Diego, CA, USA). The pooled library was then denatured and prepared for loading on an Illumina MiSeq cartridge with a 5% PhiX Control.

The sequences for forward and reverse PCR primers: Forward: AATGATACGGCGACCACCGAGATCTACAC-barcode-TATGGTAATTGTGTGCCAGCMGCCGCGGTAA; Reverse: CAAGCAGAAGACGGCATACGAGAT-barcode-AGTCAGTCAGCCGGACTACHVGGGTWTCTAAT

### METABOLOMICS

#### Mass Spectrometry Sample Preparation

Metabolomics analyses were performed by the Calgary Metabolomics Research Facility at the University of Calgary (Calgary, Alberta, Canada).

For sample preparation, saliva, aspirate, SIMBA capsule and fecal samples were collected from patients with IBS and healthy controls.

*For saliva, aspirate and SIMBA capsule samples*, 40 μL of samples were transferred to the microcentrifuge tube and homogenized by pipetting and 40 μL of pre-chilled 100% methanol was added to each sample. 120 μL of 50% pre-chilled methanol/water was added to each sample. Samples were incubated at 4°C for 30 minutes and vortexed every 10 minutes to ensure proper mixing, followed by incubation at -80°C for at least 1 hour. Samples were then centrifuged at max speed (~13,000 rpm or ~21,000 x g). 50 μL of supernatant was transferred to a clean 1.7 mL microcentrifuge tube and 150 μL of 50% pre-chilled methanol/water was added to each sample.

*For fecal samples*, 500-1000 μL of pre-chilled 50% methanol/water was added to 100-200 mg of fecal material (5 x v/w) and then homogenized by vortexing at max speed for 1 minute. After homogenization, the samples were incubated at 4°C for 30 minutes. Fecal samples were centrifuged at max speed (~13,000 rpm or ~21,000 x g) for 10 minutes at 4°C. 500 μL of supernatant were transferred to the 1.7 mL microcentrifuge tube. The samples were incubated at -80°C for at least 1 hour before the analysis.

**Semi-targeted metabolomics analysis of plasma.** Plasma samples were analyzed by liquid chromatography mass spectrometry (LC-MS) using a Q Exactive HF Hybrid Quadrupole-Orbitrap Mass Spectrometer (Thermo Fisher Scientific, Waltham, MA, USA). Chromatography was performed using a 2.1 mm x 100 mm long Synchronism HILIC (Thermo Fisher Scientific, Waltham, MA, USA) LC column packed in-house with 3 μm porous Hyperarc particles. The analytical methods used in this study have been described in detail elsewhere [1]. Briefly, metabolites were eluted with a gradient of acetonitrile with 0.1% formic acid (solvent B) transitioning into 20mM ammonium formate pH 3.0 in $H_2O$ (Solvent A). LC-MS data were acquired in negative ion mode in full scan mode with a resolution of 240,000 and a scan range of 70-1000 m/z. Maven, an open-source software, was used to process metabolomics data obtained by LC-MS [2,3].

**Targeted LC-MS quantification of bile acids.** LC-MS methods used to quantify bile acids were adapted from existing LC-MS methods [4]. Chromatography was completed using a Vanquish™ LC system coupled to a TSQ Quantum™ Access MAX triple quadrupole mass spectrometer (Thermo Fisher Scientific, Waltham, MA, USA) equipped with an electrospray ionization (HESI-II) probe. Chromatographic separation was achieved on a Hypersil GOLD TM C18 column (200 X 2.1 mm, 1.9 µm, Thermo Fisher Scientific) using a binary solvent system composed of LC-MS grade $H_2O$ containing 5 mM ammonium acetate and 0.1% (v/v) formic acid (Solvent A) and LC-MS grade methanol containing 5 mM ammonium acetate and 0.1% (v/v) formic acid (solvent B). The following 40 min gradient, with chromatographic resolution of monitored isobaric bile acids (CDCA/DCA/HDCA/UDCA and TCDCA/TDCA), was used: 0-5 min, 60% B; 5-12 min, 60-80% B; 12-20 min, 80% B; 20-24 min, 80-100% B; 24-34 min, 100% B, 34-35 min, 100-60% B, 35-40 min, 60%B. The flow rate was 250 µL min-1 and the sample injection volume 5 µL. The auto sampler was kept at 6°C and the column at 30°C. MS/MS data were acquired in positive electrospray ionization mode with the mass spectrometer operating in selected reaction monitoring (SRM) mode. Fragmentation parameters were optimized using the EZ Tune program with direct infusion of the analytical grade bile acid standards (50 µM each in 50% solvent A-solvent B). For each bile acid, a pair of quantifiers (Quan) and qualifier (Qual) ions was selected. Subsequently, the following transitions were monitored, with a scan time of 0.05 sec: Bile acid, [M+NH4]+ m/z Parent ion → m/z Quan (CE), m/z Qual (CE), where CE is the collision energy (V). CA, m/z 426.3 → m/z 355.3 (20), m/z 373.3 (14); CDCA/DCA/HDCA/UDCA, m/z 410.3 → m/z 357.3 (14), m/z 339.3 (15); CDCA-Me, m/z 508.4 → m/z 371.3 (17), m/z 339.3 (22); GCA, m/z 483.3 → m/z 412.4 (20), m/z 430.4 (17); GCDCA, m/z 467.4 → m/z 414.3 (19), m/z 432.4 (13); LCA, m/z 394.3 → m/z 359.4 (12), m/z 135.4 (35); TCA, m/z 533.3 → m/z 462.3 (22), m/z 337.3 (28); TCDCA,/TDCA m/z 517.3 → m/z 464.4 (19), m/z 482.4 (13); TLCA, m/z 501.3 → m/z 466.4 (16), m/z 341.3 (24). Electrospray ionization source conditions were as follows: spray voltage of 3000 V, vaporizer temperature of 200°C, sheath gas of 20 psi, auxiliary gas flow of 2 (arbitrary units) and sweep gas flow of 1 (arbitrary units), capillary temperature of 200°C.

Bile acids in the biological samples were quantified using external standard calibration. A freshly prepared working solution, containing CA, CDCA, DCA, HDCA, LCA and UDCA at 200 µM (final concentration), GCA, GCDCA, TCA, TCDCA, TDCA at 50 µM (final concentration), TLCA at 30 µM (final concentration) and CDCA-Me at 0.5 µM (final concentration) in methanol, was further diluted (1:2) in methanol to prepare 12 calibration solutions. Data analyses, on the converted mzXML files, were conducted in El-MAVEN [2,3] using integrated peak intensity (area under the curve). Metabolite concentrations were calculated following the Bishop method [5].

**Targeted LC-MS quantification of short-chain fatty acids (SCFAs)**. Methods for quantifying short-chain fatty acids have been described in detail elsewhere [6]. Briefly, our strategy uses stable isotope labeled standards to compute absolute concentrations of SCFAs present in extracts. Samples were collected, processed and stored at -80°C until LC-MS/MS analysis was performed. A stable isotope-labeled internal standards (IS) mix (4 µL) was added to extracted samples (40 µL) cooled on ice, followed by aniline (2 µL of a 2.4 M solution in methanol) and N-(3-Dimethylaminopropyl)-N′-ethylcarbodiimide hydrochloride (EDC, 2 µL of a 1.2 M solution in $H_2O$). For fecal samples, the composition (final concentrations) of the stable isotope-labeled internal standards (IS) mix was as follows: acetic acid-1,2-$^{13}$C2, 4 mM: propionic acid-$^{13}$C3, 1 mM; butyric acid-1,2-$^{13}$C2, 1 mM; isobutyric acid-d7, 250 µM; valeric acid-d9, 500 µM and isovaleric acid-d9, 250 µM. This IS-mix was diluted (1:200) before being added to the other sample types i.e., saliva, aspirate and capsule. Samples were left on ice for 2 hours with regular shaking. Fecal samples were diluted (1:50) with $H_2O$/methanol (50:50, v/v) prior to LC-MS/MS analysis, whereas the other samples (saliva, aspirate and capsule) were analyzed undiluted.

### DATA AND STATISTICAL ANALYSIS

For 16S sequencing, paired-end forward and reverse reads were pre-processed with Fastp (v0.23.2) [7] to remove adapters and low-quality bases removed from the 3’ ends of reads (--cut_front, -cut_right) using a sliding window mean quality filter (--cut_window_size 4, –mean_cut_quality 20). Read pairs passing quality filtering were kept for downstream merging and assembly. MultiQC (v1.15) [8] was used to evaluate per-position read quality and proportion of reads remaining post-quality filtering. The Dada2 (v1.30) [9] pipeline was implemented in R (v4.3.1) [10] to further quality-filter (maxEE 2,5), denoise, merge forward and reverse reads, and remove chimeras to generate an amplicon sequence variant (ASV) feature table. ASV taxonomic annotation was also performed using Dada2 with the RDP classifier (assignTaxonomy) with pre-formatted SILVA 16S database (v138.1) [11] and exact-matching for species-level assignments.

For the downstream calculation of unifrac beta-diversity, a phylogenetic tree was also generated for ASV sequences. Unique ASV sequences were extracted and aligned generated using mafft (v7.520, default settings) [12] which was input to fasttree (v2.1.11) (default settings) [7] for maximum-likelihood phylogenetic tree construction. Visualization of the preliminary ASV phylogenetic tree and multiple-sequence-alignment (MSA) also identified the presence of potential contaminant ASVs of host-origin. These ASVs comprised a highly divergent outlier clade, the majority of which were annotated at the family level as ‘Mitochondria’, had sequence lengths deviating from the expected 253 bp for the 16S V4 region, and limited sequence homology compared to the majority of bacterial annotated ASVs, and were therefore removed from subsequent analysis.

For metabolomics analyses, data were peak-picked using El-MAVEN V.12 [3], metabolites were identified using co-elution, m/z matching, and MS/MS fragmentation following established methods [13], quantified using external calibration curves following published methods [5]. For semi-targeted metabolomics data, non-parametric analysis of variance (Kruskal-Wallis) analysis was performed to indicate differences between samples from different groups based on the raw ion intensities. False discovery rate (FDR) was obtained after Bonferroni corrections. Student’s T-test was performed to indicate differences between samples from two groups. All statistical tests were two sided, and adjusted p values below 0.05 were considered statistically significant. The diagnostic values of the metabolites were evaluated by constructing receiver operating characteristic (ROC) curves and computing the areas under the curves (AUCs) as well as sensitivities at predefined specificities.

Final data analyses were performed using R (ver 4.3.1). Visualizations were generated with custom scripts incorporating tidyverse and ggplot2 (ver 2.0.0) [14], phyloseq (ver 1.41.1) [15], ComplexHeatmap (ver 2.16.0) [16], circlize (ver 0.4.15) [17], UpSetR (ver 1.4.0) [18], cowplot (ver 1.1.1) [19], and ggh4x (ver 0.2.8) [20] packages. The rstatix (ver 0.7.2) [21] package was used to perform Chi-squared goodness of fit test of frequency-associations between X-Ray localization vs. motility groups, the Kruskal-Wallis test for shift in dispersion of capsule sample duration by motility groups, and Mann-Whitney test (FDR corrected) for pairwise testing of targeted and untargeted metabolite concentration and spectral abundance differences, respectively, across sample types. Mann-Whitney testing was also performed to assess the longitudinal reproducibility of 16S (weighted unifrac distance) and semi-untargeted metabolomics profiles (Euclidean distance of log10 transformed spectral counts) between-sample type (capsule vs. fecal between-individual differences) and sample type (within-individual capsule X-Ray vs. endoscopy; fecal X-Ray vs. endoscopy) across visits. The vegan (ver 2.6.4) [22] package was used to perform PERMANOVA (adonis2) testing of 16S microbiome compositional differences by sample types and motility groups.
