## Supplemental Figure 1 for "CLINICAL SAMPLING OF SMALL INTESTINE LUMINAL CONTENT FOR MICROBIOME MULTI-OMICS ANALYSIS: A PERFORMANCE ANALYSIS OF THE SMALL INTESTINE MICROBIOME ASPIRATION (SIMBA) CAPSULE AND BENCHMARKING AGAINST ENDOSCOPY"

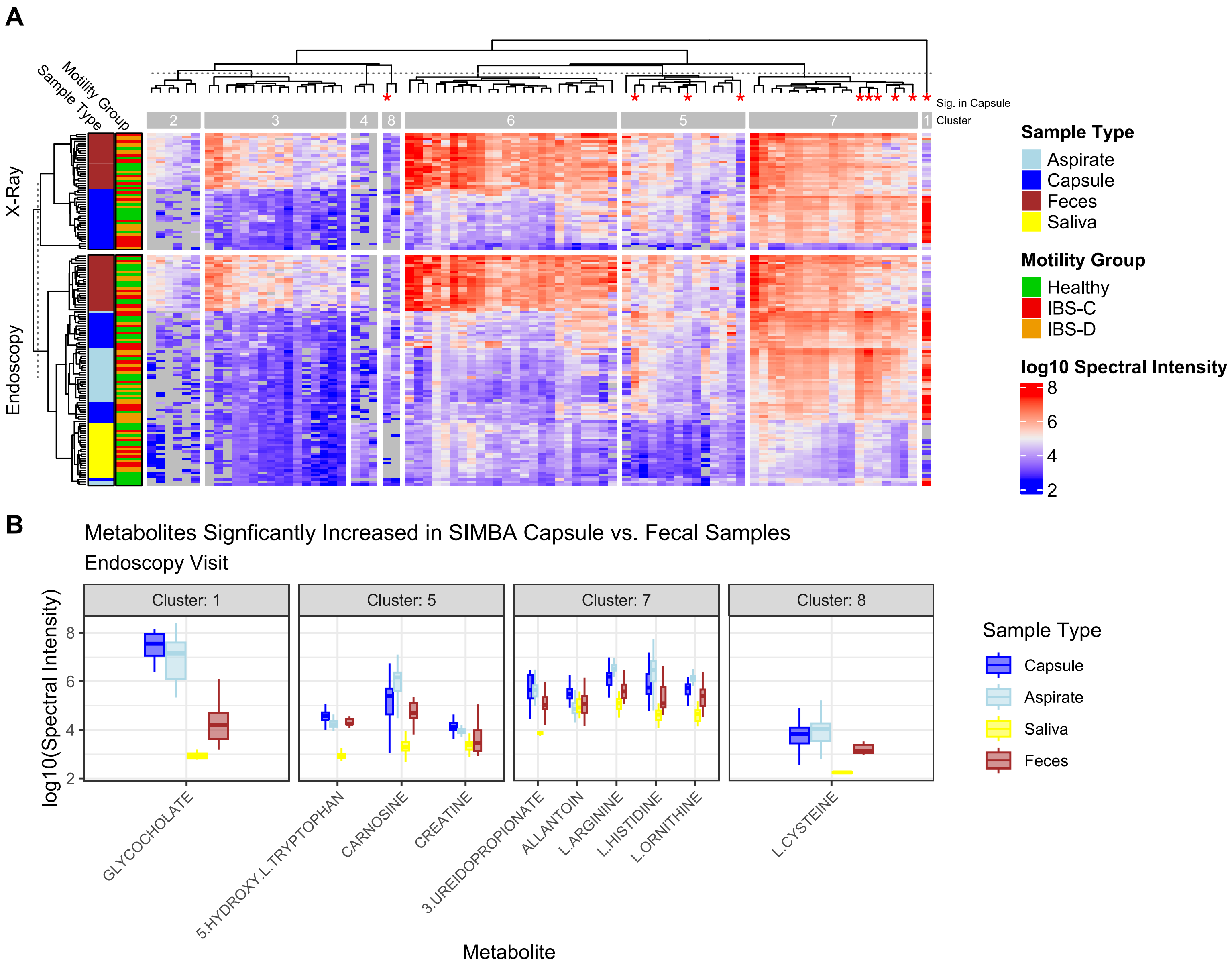


**Supplemental Figure 1: Semi-Targeted Metabolomics Profiling Across Sample Types and Visits – log10 Spectral Counts: (A)** Hierarchically clustered heatmap of log10 spectral intensities for 85 metabolites (columns) across samples (rows) collected during X-Ray (bottom) and Endoscopy visits (top). Sample dendrogram indicates distinct clustering of SIMBA capsule and fecal samples across visits, and co-clustering of SIMBA and endoscopic aspirates. The dashed-line across the column dendrogram corresponds to an optimal K-means clustering (n = 8) of metabolite profile patterns identified across SIMBA capsule/endoscopy vs. fecal samples. Red asterisks on tips of column dendrogram indicate metabolites with significantly increased log10 spectral intensities in SIMBA capsules vs. feces (Mann-Whitney test FDR adjusted p-value <= 0.05). **(B)** Boxplot of log10 spectral intensity values of metabolites significantly increased in SIMBA capsules compared to fecal samples (corresponding to asterisks indicated in panel A; Mann-Whitney FDR p-value <= 0.05).
